# Carotid Intima-media thickness, carotid distensibility and incident heart failure in older men: The British Regional Heart Study

**DOI:** 10.1101/2024.05.29.24308172

**Authors:** Atinuke Akinmolayan, A Olia Papacosta, Lucy T Lennon, Elizabeth A Ellins, Julian PJ Halcox, Peter H Whincup, S Goya Wannamethee

## Abstract

**Objectives:** Carotid intima-media thickness (CIMT) and carotid distensibility are markers of arterial change, however little is known of the prognostic value in incident HF. We aimed to assess this prospective association.

**Design:** Longitudinal analysis of data from the British Regional Heart Study, a prospective cohort study

**Participants:** 1069 men aged 71-92 years, without a diagnosis of heart failure or myocardial infarction (MI) at baseline.

**Methods:** Between 2010-2012, participants completed a questionnaire, underwent a physical examination, and provided a fasting blood sample. CIMT and carotid artery distension were measured and carotid distensibility was calculated. Cox proportional hazards modelling was used to assess the multivariate-adjusted hazard ratios of incident HF by quartiles of CIMT and distensibility.

**Results:** Adjusted for age, social class, smoking, physical activity, alcohol status, BMI, use of statins and antihypertensives, prevalent diabetes mellitus and stroke, pulse pressure, and presence of atrial arrhythmias, lower carotid distensibility was associated with increased risk of incident HF (HR 2.55, 95% CI 1.24 to 5.24, p=0.01). The association persisted after further adjustment for CIMT and incident MI (HR 2.53, 95% CI 1.23 to 5.22, p=0.01). Higher CIMT was associated with increased risk of incident HF (HR 2.20, 95% CI 1.14 to 4.23, p=0.02). However, this was attenuated after adjustment for both CIMT and incident MI (HR 1.64, 95% CI 0.84 to 3.21, p=0.1).

**Conclusions:** Low carotid artery distensibility, but not high carotid intima-media thickness (CIMT), was associated with an increased risk of incident heart failure, independent of the development of myocardial infarction.

## Introduction

In the United Kingdom, heart failure (HF) affects approximately 920,000 people with 200,000 new cases annually(1). Primarily a disease affecting older adults, at least 5% of people over the age of 75 are affected with heart failure, with prevalence increasing with age to 15% in people over 85(2). It is the only cardiovascular disease (CVD) increasing in prevalence despite advances in medical care, likely due to heart failure being a final complication for several cardiac conditions that affect cardiac contractility(3). Given the ageing of many developed country populations and increased survival from acute cardiac events, those affected by heart failure are often debilitated by multimorbidity, which in turn contributes to increased mortality, use of public health resources and healthcare cost (3). Despite advances in medical care, heart failure remains a progressive and incurable chronic disease with significant impacts on the individual, their family and society. Thus, effective preventative strategies are urgently needed.

Although the typical onset of heart failure appears later in life, evidence suggests that subclinical vascular modifications develop much earlier. Early, accurate diagnosis is critical to improving heart failure outcomes. On a structural level, heart failure is characterised by a combination of reduced contractility, ventricular remodelling, systolic and/or diastolic dysfunction, ultimately leading to the inability of the heart to meet the circulatory demands of the body. Atherosclerosis is a common cause of heart failure(4), affecting the coronary as well as carotid arteries. Development of atherosclerosis is a gradual process with a long subclinical state before it clinically manifests as cardiovascular disease and can be measured non-invasively in its early stages. Carotid intima-media thickness (CIMT) is widely reported as being an early indicator of subclinical atherosclerosis and has been shown to be associated with incident heart failure, possibly due to shared atherosclerotic pathways or through its relationship with arterial distensibility(5–7). Reduced carotid and aortic distensibility, indicators of vascular structure and function, have been shown to be early markers of subclinical vascular modification, however little is known of the prognostic value in incident CVD events(8, 9). Understanding the relationship between these markers of arterial change and incident heart failure, may eventually lead to targeted therapeutic interventions.

The contributions of these subclinical vascular measures, independent of well-established cardiovascular risk factors, on the risk of incident heart failure is not well established. This can be accounted for by conflicting reports in the literature (7, 10). In this study we postulated that non-invasive vascular measures CIMT and carotid distensibility may be related to an increased risk of incident HF independent of classical cardiovascular risk factors. We examined prospective associations between CIMT and carotid distensibility and the risk of incident heart failure in a cohort of older British men considering a wide range of cardiovascular risk factors.

## Methods

The British Regional Heart Study is a prospective study of 7735 men from 24 British towns recruited and examined at baseline between 1978 and 1980, comprising a socioeconomically and geographically representative cohort. Over 99% of participants were of White European ethnicity (11). The cohort has been followed up by a combination of primary care record reviews, NHS Central Register flagging, periodic postal questionnaires, and re-examinations. A 30-year re-examination took place in 2010-12, with all 3137 surviving men invited to attend (then aged 71-92 years). Attendees completed a questionnaire, underwent physical examination, and provided a fasting blood sample (12). At the 30-year follow up, a total of 2137 (68%) men completed the questionnaire, of whom 1722 (55%) men underwent a physical re-examination. The men were asked whether a doctor had ever told them that they had myocardial infarction (heart attack, coronary thrombosis), stroke, or diabetes and to bring their medication to the examination session. Blood samples were collected after fasting for a minimum of six hours and were stored at −70°C. Ethical approval has been obtained from all relevant local research ethics committees.

In this study, the components of the data used from the questionnaire include CVD risk factors, medications, and existing comorbidities. Blood pressure, body mass index (BMI) and presence of arrhythmia on ECG were used from the data collected at the physical examinations. NT-pro-BNP and plasma glucose levels were used from the results obtained through blood test analysis.

### CVD risk factors

CVD risk factors were assessed at the 30-year re-examination between 2010-2012. Information on the measurement and classification of alcohol intake, smoking status, physical activity, and social class assessed by questionnaire have already been previously described (13, 14). The use of antihypertensive medication was based on self-reported medication history and review of codes from the British National Formulary (BNF). Anthropometric measurements of body weight and height were used to calculate the BMI as weight/(height^2^) (kg/m^2^). Blood pressure was recorded as a mean of two measurements done in succession in the right arm using an Omron 907 blood pressure machine, with the subject seated. The values used in the analysis were adjusted for observer and cuff size differences. Pulse pressure was then calculated by subtracting the mean diastolic blood pressure measurement from the mean systolic blood pressure measurement. A 12-lead ECG was performed as part of the physical examination assessment and ECG Minnesota codes were used in algorithms to classify arrhythmias. In this study, the atrial arrhythmias namely Atrial Fibrillation (AF), Atrial Flutter and Atrial Tachycardia have been grouped together.

The study participants were asked to fast for a minimum of 6 hours prior to their follow-up physical examination for fasting blood tests to be taken. NT-pro-BNP was measured in plasma samples on an automated clinically validated immunoassay analyser (e411, Roche Diagnostics, Burgess Hill, United Kingdom) using the manufacturers’ calibrators and quality control reagents. The lower limit of sensitivity was 5 pg/ml. Plasma glucose was measured by a glucose oxidase method (15) using a Falcor 600 automated analyser. Prevalent diabetes included men with a doctor-diagnosis of diabetes and men with fasting blood glucose ≥7 mmol/L.

### Non-invasive vascular measures

Left and right carotid arteries were imaged by two experienced vascular technicians. All studies were performed using the Z.One Ultra ultrasound system (Zonare Medical Systems, Mountain View, CA) with a 5-10-mHz linear probe. Carotid artery distension and carotid artery intima-media thickness (CIMT) (the distance between the leading edge of the intima and the media-adventitia interface) were measured using the longitudinal images of the carotid artery on the Carotid Analyser Software (Medical Imaging Applications, Iowa City, IA). In the images, a 5-10mm plaque-free region of interest was selected, at least 1cm from the bifurcation. Mean CIMT was calculated from IMT measurements taken from three end-diastolic images. Maximum and minimum carotid artery diameters were taken from three consecutive waveforms, and mean distension was calculated by subtracting baseline diameter from peak diameter. Carotid distensibility coefficient was calculated using the methods described by Dijk et al., (2005) (16) as follows: ((2 x Mean Distension/Baseline diameter)/Mean PP (kPa)) * 1000. A coefficient of variation (CV) was calculated, with respect to the inter- and intra-observer reproducibility for both CIMT (inter n=109 CV=7.1%, intra n=30 CV=5.1%) and carotid distension (inter n=109 CV=9.2%, intra n=30 CV=11.9%) (17). In total 1696 men had their CIMT measured, and 1687 men had their distensibility calculated.

### Follow-up

All men were followed up to June 2018 for cardiovascular morbidity and mortality. Mortality was obtained via the National Health Service Central Register. Incident heart failure was defined as the first occurrence of a new heart failure diagnosis after the 30-year re-examination in 2010-12. Incident HF was defined as a confirmed doctor’s diagnosis of HF from primary care records, and verified, where possible, using clinical information from primary and secondary care records, as well as from death certificates with International Classification of Diseases (ICD)-9 code 428. Time to incident HF was defined as the time from the baseline re-examination in 2010-12 to the incident event. For subjects who did not develop heart failure, the end of follow up was either the date of the 2018 record review or the date of death, whichever came first.

### Statistical Analyses

In this study we hypothesised that increased CIMT and reduced carotid distensibility may be associated with an increased risk of incident HF independent of classical cardiovascular risk factors.

The men were divided into equal quartiles based on the CIMT or distensibility distribution. We report counts with percentages for categorical variables and the correlation coefficient and p-value for continuous variables. Cox proportional hazards modelling was used to assess the multivariate-adjusted hazard ratios (HR; relative risk) of incident HF by quartiles of CIMT and distensibility and per 1-SD increase in CIMT and distensibility. Person-years at risk were calculated from the date of baseline until the date of incident heart failure. A series of models were generated, adjusting for potential confounders: age, social class (manual/non-manual), smoking status (never/recent ex-smoker/long term ex-smoker/current), physical activity (inactive/active), alcohol (moderate-heavy drinker), BMI group (< 20 kg/m^2^/20– 24.9 kg/m2/25–29.9 kg/m^2^/≥30 kg/m^2^), diabetes mellitus at baseline (yes/no), history of stroke at baseline (yes/no), prevalent MI at baseline (yes/no), taking statins at baseline (yes/no), taking antihypertensives at baseline (yes/no), arrythmia at baseline (yes/no). In multivariate analyses blood markers were fitted as a continuous variable. Hazard ratios and 95% confidence intervals are shown for the analyses.

All analyses were performed using SAS version 9.4 (SAS, Cary, North Carolina)

## Results

89 men with a confirmed diagnosis of HF at baseline were excluded from analysis. Men with missing data for any one of the variables included in the Cox proportional hazard models were excluded from the models as were men with prevalent MI at baseline (n=217).

### Baseline characteristics

The men were divided into equal quartiles based on the CIMT or carotid distensibility distribution (Q0-Q3). Baseline characteristics of all men who had their mean CIMT measured is given in Table 1. Compared to men with a lower mean CIMT, men in the highest mean CIMT quartile were more likely to be of a manual social class (50.37%), physically inactive (42.07%) and be either a current or ex-smoker (66.67%). They were also more likely to have cardiovascular related comorbidities at baseline such as stroke (11.69%), diabetes mellitus (15.94%) and cardiac arrythmia (13.35%). Men in the lowest mean CIMT quartile were more likely to have a higher mean distensibility. This inverse relationship between CIMT and carotid distensibility is emphasised in the correlation matrix given in Table 2. It shows a significant negative correlation between mean CIMT and mean distensibility (r = −0.208, p<0.0001). The correlation matrix shown in Table 2 also demonstrates that mean CIMT increases with age, BMI, systolic blood pressure, pulse pressure and log-NT-pro-BNP levels.

**Table 1-.** Baseline characteristics of men by CIMT quartiles.

| Characteristic | Mean CIMT quartiles |  |  |  | p-value for the Mantel-Haenszel chi-square test |
| --- | --- | --- | --- | --- | --- |
|  | 0<br>*n*=401 | 1<br>*n*=396 | 2<br>*n*=410 | 3<br>*n*=402 |  |
| **Prevalent stroke, *n* (%)** | 30 (7.5) | 34 (8.6) | 48 (11.7) | 47 (11.7) | 0.02 |
| **Manual social class, *n* (%)** | 176 (44.1) | 174 (44.1) | 173 (42.3) | 202 (50.4) | 0.1 |
| **Never smoked, *n* (%)** | 170 (42.6) | 157 (39.9) | 150 (36.8) | 133 (33.3) | 0.005 |
| **Physically inactive, *n* (%)** | 127 (34.1) | 156 (41.2) | 159 (41) | 159 (42.1) | 0.04 |
| **Moderate-Heavy drinker, *n* (%)** | 26 (6.65) | 15 (3.84) | 20 (5) | 13 (3.3) | 0.06 |
| **Prevalent DM, *n* (%)** | 58 (14.8) | 49 (12.7) | 55 (13.7) | 62 (15.9) | 0.6 |
| **On antihypertensives, *n* (%)** | 241 (61.3) | 229 (58.6) | 249 (61.2) | 230 (57.8) | 0.5 |
| **On statins, *n* (%)** | 202 (50.4) | 194 (49) | 200 (48.8) | 190 (47.3) | 0.4 |
| **Atrial arrhythmias, *n* (%)** | 32 (8) | 46 (11.7) | 49 (12) | 53 (13.4) | 0.02 |
| **High carotid distensibility (top quartile), *n* (%)** | 113 (28.4) | 111 (27.9) | 106 (26.6) | 68 (17.1) | 0.0002 |
| Data is presented as number (percentages). DM indicates diabetes mellitus. Atrial arrhythmias include atrial fibrillation, atrial flutter or tachycardia. |  |  |  |  |  |

**Table 2-.** Mean CIMT & mean carotid distensibility correlation matrix.

|  | **Mean CIMT** | **Mean Carotid Distensibility** |
| --- | --- | --- |
| **Mean CIMT** | | $r = -0.21$<br>$p < 0.0001$ |
| **Age** | $r = 0.19$<br>$p < 0.0001$ | $r = -0.30$<br>$p < 0.0001$ |
| **BMI** | $r = 0.081$<br>$p = 0.0012$ | $r = -0.0034$<br>$p = 0.89$ |
| **Pulse pressure** | $r = 0.15$<br>$p < 0.0001$ | $r = -0.20$<br>$p < 0.0001$ |
| **Systolic BP** | $r = 0.091$<br>$p = 0.0003$ | $r = -0.32$<br>$p < 0.0001$ |
| **NT-Pro BNP (Log)** | $r = 0.089$<br>$p = 0.0006$ | $r = -0.13$<br>$p < 0.0001$ |
| BMI indicates body mass index; BP indicates blood pressure; NT-Pro BNP indicates N-terminal-pro brain natriuretic peptide |  |  |

Baseline characteristics of all men who had their mean carotid distensibility calculated is given in Table 3. Compared to men with a higher mean carotid distensibility, men in the lowest mean carotid distensibility quartile were more likely to be inactive (44.56%) and be either a current or ex-smoker (67.34%). They were also more likely to have cardiovascular related comorbidities at baseline such as stroke (11.53%) and cardiac arrhythmias (20%). As mentioned previously, there is a negative correlation between mean CIMT and mean carotid distensibility, which is reinforced by there being more men in the lowest mean carotid distensibility quartile (39.45%) falling into the highest mean CIMT quartile, compared to the highest mean carotid distensibility quartile (17.09%). Conversely to CIMT, the correlation matrix shown in Table 2 demonstrates a negative relationship between mean carotid distensibility and age, BMI, pulse pressure, systolic blood pressure and log-NT-pro-BNP.

**Table 3-.** Baseline characteristics of men by mean carotid distensibility quartiles.

| Characteristic | Mean carotid distensibility quartiles |  |  |  | p-value for the Mantel-Haenszel chi-square test |
| --- | --- | --- | --- | --- | --- |
|  | 0<br>*n*=399 | 1<br>*n*=400 | 2<br>*n*=400 | 3<br>*n*=399 |  |
| **Prevalent stroke, *n* (%)** | 46 (11.5) | 37 (9.3) | 35 (8.8) | 37 (9.3) | 0.3 |
| **Manual social class, *n* (%)** | 178 (44.7) | 184 (46.5) | 183 (45.8) | 177 (44.4) | 0.9 |
| **Never smoked, *n* (%)** | 130 (32.7) | 157 (39.4) | 149 (37.5) | 169 (42.8) | 0.009 |
| **Physically inactive, *n* (%)** | 168 (44.6) | 150 (39.4) | 147 (38.8) | 132 (35.4) | 0.01 |
| **Moderate-Heavy drinker, *n* (%)** | 21 (5.4) | 20 (5.1) | 18 (1.2) | 16 (1) | 0.4 |
| **Prevalent DM, *n* (%)** | 58 (14.8) | 43 (11) | 68 (17.4) | 54 (14.1) | 0.6 |
| **On antihypertensives, *n* (%)** | 240 (60.8) | 232 (58.4) | 237 (60.2) | 234 (59.7) | 0.9 |
| **On statins, *n* (%)** | 187 (46.9) | 196 (49) | 199 (49.8) | 196 (49.1) | 0.5 |
| **Atrial arrhythmias, *n* (%)** | 79 (20) | 39 (9.8) | 39 (9.8) | 22 (5.6) | <0.0001 |
| **High CIMT (top quartile), *n* (%)** | 157 (39.5) | 98 (24.5) | 75 (18.8) | 68 (17.1) | <0.0001 |
| Data is presented as number (percentages). DM indicates diabetes mellitus. Atrial arrhythmias include atrial fibrillation, atrial flutter, or tachycardia. |  |  |  |  |  |

### CIMT & incident HF risk

Mean follow-up time was 6.04 years (SD 1.85 years). Table 4 shows the rate of incident HF (per 1000 person-years) by quartile of the CIMT distribution. The lowest mean CIMT quartile (Q0) was used as a reference group. The rate of incident HF increases from 6.58 per 1000 person-years in Q0 to 19.7 per 1000 person-years in Q3.

**Table 4-.** Adjusted hazard ratios (95% CI) for incident heart failure per CIMT quartile.

|  | Quartiles of mean CIMT (mm) |  |  |  |  |
| --- | --- | --- | --- | --- | --- |
|  | Q0<br>< 0.70 | Q1<br>0.70 to 0.78 | Q2<br>0.78 to 0.90 | Q3<br>0.90 to 1.79 |  |
| Total number of men | 338 | 359 | 348 | 350 |  |
| Number of HF events | 14 | 21 | 17 | 39 |  |
| Rate/1000 pyrs | 6.58 | 9.27 | 8 | 19.1 |  |
| **Models** |  |  |  |  | **Per unit SD increase** |
| Model 1, HR (95% CI) | 1.0 (ref.) | 1.41 (0.72, 2.77) | 1.06 (0.52, 2.16) | 2.40 (1.29, 4.46)** | 1.36 (1.15, 1.62) |
| Model 2, HR (95% CI) | 1.0 (ref.) | 1.30 (0.65, 2.60) | 0.93 (0.44, 1.96) | 2.11 (1.10, 4.07)** | 1.32 (1.10, 1.59) |
| Model 3, HR (95% CI) | 1.0 (ref.) | 1.29 (0.64, 2.61) | 0.97 (0.46, 2.05) | 2.21 (1.15, 4.26)** | 1.35 (1.12, 1.63) |
| Model 4, HR (95% CI) | 1.0 (ref.) | 1.30 (0.64, 2.63) | 0.94 (0.44, 2.00) | 2.20 (1.14, 4.23)** | 1.35 (1.12, 1.63) |
| Model 5, HR (95% CI) | 1.0 (ref.) | 1.28 (0.63, 2.59) | 0.95 (0.45, 2.01) | 1.93 (0.99, 3.75) | 1.30 (1.07, 1.58) |
| **Final model variations** |  |  |  |  |  |
| Model 5a, HR (95% CI) | 1.0 (ref.) | 1.15 (0.57, 2.34) | 0.79 (0.37, 1.70) | 1.64 (0.84, 3.21) | 1.27 (1.04, 1.55) |
| * CIMT SD = 0.157. ** P < 0.05.<br>Model 1: adjusted for age. Model 2: adjusted for age, social class, smoking, physical activity, alcohol intake, BMI, statin medication, prevalent diabetes, prevalent stroke. Model 3 : adjusted for covariates in model 2 and systolic blood pressure and hypertensive medication. Model 4: adjusted for covariates in model 3 and arrhythmias. Model 5: adjusted for covariates in model 4 and carotid distensibility. Model 5a: adjusted for covariates in model 5 and time-dependent incident MI. |  |  |  |  |  |

Men in the highest CIMT quartile (Q3) showed a statistically significant association with incident HF risk in the age-adjusted model (HR 2.40, 95% CI 1.29 to 4.46, p=0.006) as shown in Model 1 in Table 4. The significant association was maintained after adjusting for social class, smoking, physical activity, alcohol status, BMI, use of statins, prevalent diabetes mellitus, prevalent stroke (Model 2 HR 2.11, 95% CI 1.10 to 4.07, p=0.02), systolic blood pressure and use of antihypertensives (Model 3 HR 2.21, 95% CI 1.15 to 4.26, p=0.02) and after adjustment for atrial arrhythmias (Model 4 HR 2.20, 95% CI 1.14 to 4.23, p=0.02). Adjustment for mean carotid distensibility (Model 5) attenuated the association, resulting in an association that was not significant between mean CIMT and risk of incident HF (HR 1.93, 95% CI 0.99 to 3.75, p=0.05). Subsequent adjustment of the final model for time-dependent incident MI, further attenuated the association between mean CIMT and incident HF risk in men in the highest CIMT quartile (HR 1.64, 95% CI 0.84 to 3.21, p=0.15).

Adding log-NT-pro-BNP to the fully adjusted model slightly weakened the association between incident HF mean CIMT in men in the highest quartile (HR 1.87, 95% CI 0.94 to 3.72, p=0.07).

### Carotid arterial distensibility & incident HF risk

Table 5 shows the rate of incident HF (per 1000 person-years by carotid distensibility quartile (Q0-Q3). The highest distensibility quartile (Q3) was used as a reference group. As mean carotid distensibility decreases, the rate of incident HF increases from 6.58 per 1000 person-years in Q3 to 19.77 per 1000 person-years in Q0.

**Table 5-.** Adjusted hazard ratios (95% CI) for incident heart failure per distensibility quartile.

|  | **Carotid distensibility quartiles ( x10<sup>-3</sup> kPa-1)** |  |  |  |  |
| --- | --- | --- | --- | --- | --- |
|  | **Q0**<br>< 9.25 | **Q1**<br>9.25 to < 11.75 | **Q2**<br>11.75 to 14.75 | **Q3**<br>>14.76 |  |
| Total number of men | 353 | 355 | 345 | 333 |  |
| Number of HF events | 40 | 20 | 16 | 14 |  |
| Rate/1000 pyrs | 19.77 | 9.21 | 7.3 | 6.58 |  |
| **Models** |  |  |  |  | **Per unit SD increase** |
| Model 1, HR (95% CI) | 2.26 (1.20, 4.25)** | 1.22 (0.61, 2.42) | 1.01 (0.49, 2.06) | 1.0 (ref.) | 0.74 (0.58, 0.94) |
| Model 2, HR (95% CI) | 2.46 (1.22, 4.98)** | 1.44 (0.68, 3.05) | 1.14 (0.52, 2.50) | 1.0 (ref.) | 0.73 (0.57, 0.95) |
| Model 3, HR (95% CI) | 2.72 (1.33, 5.55)** | 1.51 (0.71, 3.20) | 1.18 (0.54, 2.59) | 1.0 (ref.) | 0.70 (0.54, 0.91) |
| Model 4, HR (95% CI) | 2.55 (1.24, 5.24)** | 1.45 (0.69, 3.09) | 1.13 (0.51, 2.49) | 1.0 (ref.) | 0.71 (0.55, 0.93) |
| Model 5, HR (95% CI) | 2.31 (1.12, 4.77)** | 1.39 (0.66, 2.96) | 1.18 (0.53, 2.59) | 1.0 (ref.) | 0.76 (0.58, 0.99) |
| **Final model variations** |  |  |  |  |  |
| Model 5a, HR (95% CI) | 2.53 (1.23, 5.22)** | 1.57 (0.73, 3.38) | 1.32 (0.60, 2.91) | 1.0 (ref.) | 0.73 (0.57, 0.95) |
| * Carotid distensibility SD = 4.177. ** P < 0.05.<br>Model 1: adjusted for age. Model 2: adjusted for age, social class, smoking, physical activity, alcohol intake, BMI, statin medication, prevalent diabetes, prevalent stroke. Model 3 : adjusted for covariates in model 2 and pulse pressure and hypertensive medication. Model 4: adjusted for covariates in model 3 and atrial arrhythmias. Model 5: adjusted for covariates in model 4 and CIMT. Model 5a: adjusted for covariates in model 5 and time-dependent incident MI. |  |  |  |  |  |

Men in the lowest carotid distensibility quartile (Q0) showed significantly increased risk of incident HF compared to the men in the highest quartile (Q3) in the age-adjusted model (HR 2.26 95% CI 1.20 to 4.25, p=0.01) (Table 5). This significant association was maintained after adjusting for social class, smoking, physical activity, alcohol status, BMI, use of statins, prevalent diabetes mellitus, prevalent stroke (Model 2 HR 2.46, 95% CI 1.22 to 4.98, p=0.01) and after adjusting for pulse pressure and use of antihypertensives (Model 3 HR 2.72, 95% CI 1.33 to 5.55, p=0.006). Further adjustment for presence of atrial arrhythmias made minor differences to the findings (Model 4 HR 2.55, 95% CI 1.24 to 5.24, p=0.01). The association persisted after further adjustment for CIMT (Model 5 HR 2.31, 95% CI 1.12 to 4.77, p=0.02). Subsequent adjustment of the final model for time-dependent incident MI strengthened the significant association for Q0 (Model 5a HR 2.53, 95% CI 1.23 to 5.22, p= 0.01).

Adding log-NT-pro-BNP to the fully adjusted model strengthened the association between incident HF and mean carotid distensibility in men in Q0 (HR 3.08, 95% CI 1.34 to 7.05, p=0.008).

## Discussion

### Summary

The objective of this study was to investigate the association between non-invasive vascular measures CIMT and carotid distensibility – and incident HF risk. In this cohort of British men aged between 71-92, we found both higher CIMT and lower carotid artery distensibility were significantly associated with incident HF risk, which persisted after adjustment for well-established cardiovascular risk factors and comorbidities. The significant association between these vascular measures and incident HF were found to be independent of each other, demonstrated by the models adjusting for the alternative vascular measure (Table 4 Model 5, Table 5 Model 5). Use of either parameter may improve identification of patients at risk of developing HF.

CIMT is a well validated measure of subclinical atherosclerosis (18). In this study we showed that increased CIMT (between 0.9 – 1.79mm) and therefore larger preclinical atherosclerotic lesions are associated with an increased risk of incident HF. These findings are consistent with that described by Effoe et al (5), who reported a significant association between CIMT and incident HF in a cohort of 13590 American subjects. Our results were significant despite exclusion of men with prevalent MI at baseline and after adjustment for cardiovascular risk factors, comorbidities, and related medications. It has been debated in the literature that CIMT may be associated with HF through a unique mechanism different from that causing myocardial ischaemia and infarction (5). Our study shows that this may be through changes in vascular distensibility as the association between CIMT and incident HF for Q3 becomes insignificant in Model 5 (Table 4) after adjustment for carotid distensibility. It is possible that increased CIMT is associated with structural vascular changes that impact arterial distensibility and therefore results in an increased systemic peripheral resistance, increased pressure afterload and eventually diastolic dysfunction leading to heart failure (19, 20).

Distensibility is an established marker of vascular function & structure (8). In this study, we showed that reduced carotid distensibility (less than 9.2 x 10^− 3^ kPa^− 1^) is associated with an increased risk of HF. Our findings were significant despite exclusion of men with prevalent MI at baseline and after adjustment for cardiovascular risk factors, comorbidities including time dependent incident MI, related medications as well as adjustment for CIMT. Our findings contradict previously published work by Redheuil et al, who failed to find a significant relationship between aortic distensibility and incident HF in a cohort of 3675 American participants (8). It has been reported that a reduction in vascular distensibility has several consequences on the cardiovascular system, which may in turn contribute to the development of heart failure (21). These consequences include increased left ventricular afterload, increased systolic BP and pulse pressure, reduced diastolic BP leading to reduced cardiac perfusion and enhanced intravascular pressure resulting in premature atherosclerotic development (8, 21).

NT-pro-BNP is a biomarker strongly associated with HF. Its’ synthesis predominately occurs in ventricular myocytes in response to myocyte stretch and locally in the area surrounding a myocardial infarction (22). Despite the widely reported relationship between NT-pro-BNP and HF and the correlation shown in our study between NT-pro-BNP, CIMT, and carotid distensibility (Table 2), when adjusted for in the models, NT-pro-BNP weakened the association between CIMT and incident HF and strengthened the association between carotid distensibility and incident HF. This again suggests that the development of HF may be better explained by changes in carotid distensibility and also signifies that to better understand the mechanisms that could contribute to developing HF, it may be necessary in future studies to differentiate between the types of HF developed – for example HF with preserved ejection fraction (HFpEF) and HF with reduced ejection fraction (HFrEF) as the pathological pathways may differ.

### Strengths & limitations

To our knowledge, our study is the first of its kind to study the relationship between CIMT and distensibility and risk of incident HF in an older British male cohort. Our study has a relatively long follow-up, with a detailed baseline examination which allows for adjustment for potential confounders. Our study also investigated two different types of vascular measures, which are markers of slightly different components of vascular ageing. These measures are non-invasive, reliable, low-cost, and easy to measure, which makes it simple to replicate in research and in clinical practice.

Excluding men with prevalent MI at baseline was key in our study because assessing the value of subclinical markers of CVD is of limited use in people with clinically apparent CVD, particularly as there is a distinct increased risk of HF in people with a history of MI. This point is reinforced by the attenuation of the association between CIMT and incident HF in the model variation 5a (Table 4) which adjusts for time-dependent incident MI in the men included in the cohort. This attenuation was seen in the models assessing CIMT and incident HF but not in the models assessing carotid distensibility and incident HF.

Our definition of incident HF relies on physician-diagnosed HF, which may underestimate the true HF incidence in the population. We also did not collect data on HF subtype and therefore we were unable to determine the type of HF more likely to be developed in association with CIMT and carotid distensibility. As mentioned, it may be that although the different HF subtypes may overlap with their presenting clinical features, they may have different underlying pathophysiological mechanisms. Although our cohort was socioeconomically and geographically representative of Britain, it consisted of males and was almost entirely of White ethnic origin, which may limit the generalisability of the findings to women and people from other ethnic groups. Effoe et al conducted a similar study assessing the relationship between CIMT and incident HF in 13 590 male and female participants who self-reported as either White or Black (5). They found that results were similar across race and gender groups (5).

### Implications for future research

CIMT and carotid distensibility have been shown to be reliable and low-cost measures of vascular function. Both are easily measured non-invasively in a clinical setting with ultrasound. Further longitudinal studies should report on the associations between these vascular measures and other CVD events such as incident stroke, MI, and mortality. It would also be of value to determine whether there is a clinically significant threshold for raised CIMT and reduced carotid distensibility and whether these are possible therapeutic targets. Future studies could also look to incorporate these measures into a prediction tool for HF risk.

## Data Availability

Data are available from the British Regional Heart Study subject to request approval: https://www.ucl.ac.uk/epidemiology-health-care/research/primary-care-and-population-health/research/ageing/british-regional-heart-study-brhs/brhs-2

## Acknowledgments

Author contributions, using CRediT taxonomy:

Atinuke Akinmolayan: Conceptualisation, methodology, software, formal analysis, writing - original draft

A Olia Papacosta: Software, formal analysis, investigation

Lucy T Lennon: Investigation, data curation, project administration

Elizabeth A Ellins: Methodology, investigation, writing-review and editing

Julian PJ Halcox: Methodology, investigation, writing – review and editing

Peter H Whincup: Writing – review and editing, funding acquisition

S Goya Wannamethee: Writing – review and editing, formal analysis, methodology, supervision, funding acquisition

This work uses data provided by patients and collected by the NHS as part of their care and support.

## Abbreviations

HF: heart failure
CIMT: carotid intima-media thickness

